# Detection of diphtheria toxin-producing *Corynebacterium ramonii* in wounds of an urban, inner-city population in Vancouver, Canada, 2019-2023

**DOI:** 10.1101/2024.09.26.24314455

**Authors:** Christopher F. Lowe, Gordon Ritchie, Chiara Crestani, Miguel Imperial, Nancy Matic, Michael Payne, Aleksandra Stefanovic, Diana Whellams, Sylvain Brisse, Marc G. Romney

## Abstract

We conducted a retrospective chart review and whole-genome sequencing of specimens with presumptive *Corynebacterium ulcerans* in Vancouver from July 2019 to July 2023. WGS confirmed 8/14 wound culture isolates as *C. ramonii*. Two distinct clusters of *C. ramonii* were identified with clinical presentations similar to cutaneous *C. diphtheriae*.

**Article Summary:** Two distinct clusters of C. ramonii were identified in Vancouver, BC with clinical presentations similar to cutaneous C. diphtheriae.

## Introduction

*Corynebacterium ramonii*, a member of the *Corynebacterium diphtheriae* species complex, has recently been differentiated from *Corynebacterium ulcerans*(1). In addition to genomic differences, there may be epidemiological differences: *C. ulcerans* is associated with zoonotic transmission (predominantly infections in cats and dogs), while *C. ramonii* is suspected of potential human-to-human transmission, as its zoonotic character has not been established(1). *C. ulcerans* can present with respiratory or cutaneous infection similar to *C. diphtheriae*(2,3). To date, there have been 14 isolates characterized as *C. ramonii* in the literature(1), some of which have been reported to harbour the diphtheria toxin gene, potentially underscoring the clinical and public health implications of correctly identifying this organism.

In Canada, where the incidence of *C. ulcerans* is very low, there was a 35% increase in toxin testing referrals to the National Microbiology Laboratory (NML, Winnipeg, MB) between 2006–2019; twenty-two cases of *C. ulcerans* infection were identified (45% diphtheria toxin positive)(4). While there have been multiple reports of non-toxigenic cutaneous diphtheria (*C. diphtheriae*) in the inner-city of Vancouver, there have only been sporadic reports of toxigenic *C. ulcerans*(5,6). In response to the detection of additional *C. ulcerans* cases in this community – most of which were subsequently identified as *C. ramonii* – we conducted a clinical, epidemiological, and genomic review of infections caused by this novel species.

## Methods

A retrospective review of all specimens with presumptive “*C. ulcerans*” (based on MALDI-ToF) was performed by the St. Paul’s Hospital (SPH) microbiology laboratory and LifeLabs (Vancouver, BC) from January 2019 to July 2023. Chart extraction using electronic medical records (EMR) was conducted for patients seen at SPH (no clinical information available for LifeLabs’ cases) including: hospital course (inpatient/critical care admission); antibiotics; 30-day mortality; and risk factors for infection (housing status, substance use, livestock/domestic animal exposure). Bacterial cultures with *C. ulcerans* were characterized utilizing the MALDI Biotyper® Sirius System (Bruker). At SPH, antibiotic gradient strips (Etest, bioMérieux) were used to perform antibiotic susceptibility testing for penicillin, erythromycin, clindamycin and vancomycin(7). Toxin testing (PCR) and modified Elek test were performed at NML. For whole-genome sequencing (WGS), isolated bacterial colonies were extracted on the MagNA Pure 24 (Roche Diagnostics) and sequenced on the GridION (Oxford Nanopore Technologies) with R10.4.1 flowcells. Data acquisitioning and base-calling into fast5 files was performed on MinKNOW (v.23.07.12) and Guppy (v.6.4.6). Raw FASTQ files were assembled with Flye v2.9. Genome assembly quality was checked with QUAST v5.2.0 and analysed with diphtOscan(8–10). A maximum-likelihood phylogeny with IQ-tree v2.3.4 and a GTR+G model was inferred from a core gene alignment obtained with panaroo v1.5.0 and visualised with Microreact [https://microreact.org/project/rgLWfs1derHFm4K5ShbxgJ-c-ramonii-and-c-ulcerans-vancouver-canada](11–13). Core genome multilocus sequence type (cgMLST) profiles were attributed by tagging the genomes for known alleles within BIGSdb-Pasteur [https://bigsdb.pasteur.fr/diphtheria] with the cgMLST_ulcerans scheme. Minimum-spanning trees were obtained with GrapeTree directly from the BIGSdb-Pasteur plug-in(14,15).

Research ethics approval was obtained from the University of British Columbia/Providence Health Care Research Institute (H22-03695).

## Results

There were 14 patients with cultures initially identified as *C. ulcerans* by MALDI-ToF. Further analysis of the spectra revealed 8/14 samples with a peak at 5405.40 m/z (range: 5404-5406 m/z) which has been associated with *C. ramonii*(1). WGS and genotyping confirmed these isolates as *C. ramonii*, including four sequence type (ST) 335 and four ST341 (Figure 1A), all originating from the inner-city of Vancouver (Figure 1B). Results of cgMLST typing suggest the existence of two *C. ramonii* clusters of recent transmission, with a maximum of one allelic mismatch for Cluster X and no allelic mismatches for Cluster Y (Figure 1A). The remaining six isolates were confirmed as *C. ulcerans* (ST325, ST339, ST669 [3 isolates] and ST895, a novel ST) (Figure 1C). In contrast to *C. ramonii*, they did not originate from a specific neighbourhood.

**Figure 1.**
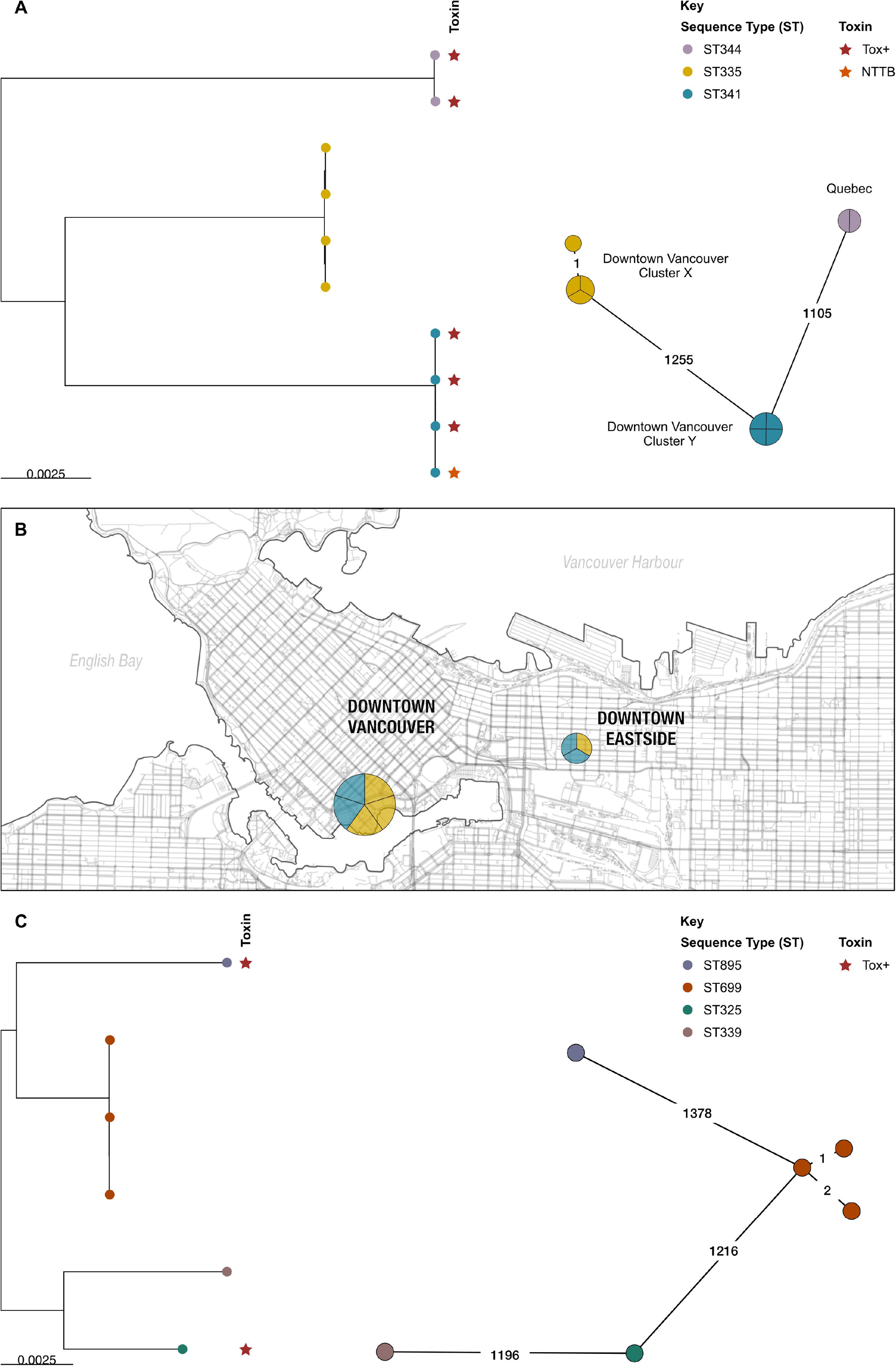
*Corynebacterium ramonii* and *Corynebacterium ulcerans* isolates from Canada. A) Maximum-likelihood phylogeny and Minimum spanning tree of eight *C. ramonii* isolates from this study (British Columbia), together with two public isolates from Canada (Quebec); two clusters of infections can be observed, corresponding to ST335 and ST341 isolates. B) Spatial map of *C. ramonii* isolates from this study, all originating from downtown Vancouver. C) Maximum-likelihood phylogeny and Minimum spanning tree of six *C. ulcerans* isolates from this study (British Columbia). Trees were rooted at midpoint.

All specimens were polymicrobial and recovered from lower extremity wounds (Table 1).

Antimicrobial susceptibility and toxigenicity data are reported in Table 1. Chart review of patients with *C. ramonii* infection revealed 3/8 required admission to hospital (average length of stay, 6 days) with no critical care admissions and no 30-day mortality. Of the 6 patients with EMR documentation, all were treated with a course of antibiotics (piperacillin-tazobactam, amoxicillin-clavulanate, trimethoprim-sulfamethoxazole, cephalexin, cefazolin or ceftriaxone). All eight patients were associated with the downtown core of Vancouver (Figure 1B), including five persons experiencing homelessness (PEH); contrastingly, *C. ulcerans* cases were distributed throughout the city outside the downtown core (based on postal code). All *C. ramonii* patients reported a history of substance use disorder. Importantly, none had documented livestock or domestic animal exposure (i.e., pets).

**Table 1.** Antimicrobial susceptibility testing, co-isolated bacteria from wound cultures and toxin testing results from *Corynebacterium ulcerans* and *Corynebacterium ramonii* isolates.

|  | *Corynebacterium ulcerans*<br>(n=6) | *Corynebacterium ramonii*<br>(n=8) |
| --- | --- | --- |
| **Antimicrobial Susceptibility** |  |  |
| Penicillin | N/A | Intermediate (8) |
| Erythromycin | N/A | Susceptible (8) |
| Clindamycin | N/A | Susceptible (1); Intermediate (7) |
| Vancomycin | N/A | Susceptible (8) |
| **Frequency of bacteria co-isolated from wound cultures** | *Enterobacterales* (4)<br>MSSA<sup>#</sup> (3)<br>Coagulase-negative<br>Staphylococci<sup>^</sup> (2)<br>β-hemolytic *Streptococcus*<sup>+</sup><br>(2)<br>*Acinetobacter baumannii*<br>(1) | $\beta$ -hemolytic *Streptococcus* * (6)<br>*Arcanobacterium haemolyticum*<br>(4)<br>MSSA<sup>#</sup> (4)<br>*Enterobacterales* (3)<br>Yeast (1)<br>*Pasteurella multocida* (1) |
| **Toxin Testing** |  |  |
| Non-toxicogenic | 4 (66.6%) | 4 (50%) |
| Non-toxicogenic *tox* gene-bearing (NTTB) | 0 | 1 (12.5%) |
| Toxigenic | 2 (33.3%) | 3 (37.5%) |

\* *Streptococcus pyogenes* and *Streptococcus dysgalactiae* subsp. *equisimilis*

## Discussion

A review of *C. ramonii* and *C. ulcerans* recovered from wound cultures in Vancouver revealed distinct epidemiological differences between these closely related organisms. *C. ramonii* cases clustered exclusively in a single geographic region (inner-city downtown Vancouver), whereas *C. ulcerans* cases resided outside of the city’s downtown core. Human-to-human transmission of *C. ramonii* has been hypothesized(1), and based on molecular, microbiological and epidemiological findings, this study suggests possible human-to-human transmission. WGS showed two distinct *C. ramonii* clusters (ST335 and ST341) within the same community. Vancouver’s downtown core has high rates of poverty, PEH and substance use, and may be conducive to transmission via close contact, such as a previous cluster of cutaneous *C. diphtheriae* associated with a single strain(5). A similar pattern appears to be emerging for patients with *C. ramonii*, all of whom are from the same community. There were no reports of animal exposure, in contrast to the association of *C. ulcerans* with zoonotic transmission. *C. ramonii* was identified with other bacteria associated with human reservoirs, either exclusively or primarily: Group A *Streptococcus*, Group G *Streptococcus, A. haemolyticum*. All wound cultures were polymicrobial in nature (and one specimen harboured *P. multocida*). Based on our preliminary review, the clinical presentation of *C. ramonii* infections aligns more closely to the presentation of cutaneous diphtheria due to *C. diphtheriae*, rather than *C. ulcerans*(5).

Our study highlights the role for MALDI-ToF identification of *C. ramonii*. The initial description of *C. ramonii* reported a unique peak at 5405.40 m/z(1). In our study, initial *C. ramonii* identification required WGS, and retrospective review of the spectra confirmed the presence of the peak, though exhibiting a broader m/z range. With MALDI-ToF widely used in clinical laboratories, the prevalence of *C. ramonii* can be more accurately estimated when peak analysis of mass spectrograms is performed to avoid misidentification as *C. ulcerans*. This also reinforces the need to continually update the MALDI-ToF spectral databases thus enabling accurate detection of emerging pathogens, such as *C. ramonii*.

Limitations of this study include the relatively small number of cases. Since wound cultures were polymicrobial, it is unclear to which degree *C. ramonii* contributed to the wound infection. No toxigenic systemic signs were observed in the three patients with diphtheria toxin-producing *C. ramonii*. In addition, we could not perform a formal case-control study, as the patients with *C. ulcerans* infection were primarily seen in outpatient clinics outside of our healthcare network, and complete medical records were not accessible.

Our findings correlate with the initial clinical description of *C. ramonii*, which appears to present similarly to cutaneous *C. diphtheriae* infections. As laboratories become increasingly proficient in differentiating *C. ulcerans* from *C. ramonii*, additional studies are required to better characterize this novel member of the *C. diphtheriae* species complex.

## Acknowledgements

We would like to thank Jennifer Bilawka and Leah Gowland for their technical support at the St. Paul’s Hospital Microbiology Laboratory. We would like to thank the BCCDC Public Health Laboratory and National Microbiology Laboratory for coordinating and performing the toxin testing for this study.

## Author biography

Dr. Lowe is a Medical Microbiologist and Infection Prevention and Control Physician at St. Paul’s Hospital in Vancouver, BC and a Clinical Professor at the University of British Columbia.

## Data availability

Sequence data for this project is available at https://bigsdb.pasteur.fr/diphtheria/, project number 30. Phylogenetic analyses *C. ulcerans* and *C. ramonii*, together with all the relevant metadata, is available in Microreact at the project URL: https://microreact.org/project/rgLWfs1derHFm4K5ShbxgJ-c-ramonii-and-c-ulcerans-vancouver-canada.

## Conflict of Interest

No relevant conflict of interests to declare.

## Funding and Open access license statement

This research was funded, in whole or in part, by Institut Pasteur. For the purpose of open access, the authors have applied a CC-BY public copyright license to any Author Manuscript version arising from this submission.

## Authorship

Conceptualization, SB, MGR; Data curation, CFL, GR, CC, MF, MI, NM, MP, AS, DW; Formal analysis, CFL, GR, CC, MGR; Sequencing and analysis, GR, CC; Funding acquisition, N/A; Investigation, CFL, GR, CC, MF, MI, NM, MP, AS, DW; Methodology, CFL, GR, CC, MF, SB, MGR; Project administration, CFL, SB, MGR; Supervision, SB, MGR; Visualization, CFL, GR, CC; Writing – original draft, CFL; Writing – review & editing, CFL, GR, CC, MF, MI, NM, MP, AS, DW, SB, MGR.

## Notes

### Competing Interest Statement

The authors have declared no competing interest.

